# CERVEX: A Foundation-Model Framework With Built-In Explainable AI for Automated Cervical Cytology Classification From Pap Smear Images

**DOI:** 10.64898/2026.09.03.26362210

**Authors:** Naphudon Sriratana, Patthadon Pongsiripreeda, Panuwitch Uasawaengboon, Napatkrit Asavarojpanich, Lojrutai Jocknoi

## Abstract

Cervical cancer is largely preventable when abnormal cells are detected early. However, manual Pap-smear screening remains labor-intensive because a specimen may contain thousands of cells, borderline abnormalities are subtle, and expert cytology review is not equally accessible. Although artificial intelligence can reduce this workload, conventional classifiers often return a label without directly exposing the evidence used and may exploit staining or image-acquisition shortcuts. To address these limitations, we present CERVEX, a pathology-foundation-model framework with an additive class-evidence readout. Each class score is computed as the spatial mean of its class-specific evidence map plus a learned bias; therefore, the heatmap and numerical score are generated by the same forward calculation. CERVEX also produces a numerical research report containing class probabilities, evidence strength, ranked candidate regions, and segmentation-derived morphology, including N:C ratio and nuclear geometry. Under repeated slide-grouped evaluation on RIVA, with labelled target-cohort cells included during training, CERVEX distinguished abnormal from normal cells with AUROC 0.911 (SD 0.009), sensitivity 0.869, and specificity 0.796. Furthermore, fixed-grid analysis without cell coordinates distinguished low-from high-grade disease across 101 graded abnormal slides with AUROC 0.823 [0.728, 0.911]. On SIPaKMeD, nucleus and complete-cell segmentation reached Dice 0.932 and 0.942, while nuclear area fraction and N:C ratio reached measurement correlations of 0.977 and 0.962, respectively. Moreover, CERVEX produced class-specific spatial evidence while a separate held-out CRIC comparison did not resolve an accuracy difference from the conventional classifier, demonstrating computation-linked explainability with only 1.9% measured runtime overhead. Ultimately, CERVEX enables automated analysis of cervical cytology microscope fields by linking cell classification, computation-linked spatial evidence, and interpretable numerical morphology indices in one pipeline. This supports transparent candidate-region review and establishes a practical foundation for future whole-slide screening and prospective clinical validation.

## I. Introduction

Cervical cancer is the fourth most common cancer in women worldwide. The World Health Organization estimated 660,000 new cases and 350,000 deaths in 2022, with the largest burden in low- and middle-income countries [1]. Persistent high-risk human papillomavirus infection causes most cases, and its slow progression makes early cytological screening effective. Cytologists report cellular abnormalities under the Bethesda System rather than assigning a tumour diagnosis [2].

Manual review creates a major screening bottleneck. Across 16 studies and 61,099 women, conventional cytology detected CIN2+ with 65.9% sensitivity (95% CI 54.9–75.3) at 96.3% specificity, compared with 92.6% sensitivity for HPV testing [3]. Thus, conventional cytology misses roughly one-third of high-grade lesions during a single visit. Cytologists also show moderate reproducibility. In the ASCUS-LSIL Triage Study, interobserver agreement on cytological interpretation reached only κ = 0.46 (95% CI 0.44–0.48), no better than punch-biopsy histology [4]. A smear may contain thousands of cells, and cytologists must assign one category despite subtle and ambiguous borderline abnormalities. This workload has motivated computer-aided screening. In a nine-hospital study, the model alone reached 0.832 accuracy for CIN2+; a separate reader-assistance experiment reduced interpretation time [5].

Deep networks now perform strongly on cervical-cell classification [6]–[9]. However, clinical adoption has lagged. When a network reports a Bethesda category without showing its evidence, the accountable pathologist cannot verify the reasoning. Moreover, the network may rely on staining, scanner, or preparation differences instead of cell morphology [10], [11]. Accuracy alone cannot distinguish these strategies.

Most explainable-AI systems estimate a saliency map after training. In contrast, classical CAM makes a class-evidence map an exact additive term in the class logit [12]. However, exact arithmetic alone cannot establish anatomical or cytological relevance, so the system must also test localization and morphology.

We developed CERVEX as a new integrated architecture for automated cervical cytology field analysis (Figs. 1, 2). CERVEX combines a pathology foundation backbone, an additive class-evidence readout, fixed-grid candidate search, contrastive evidence maps, and contour-derived morphology concepts. The architecture classifies cells, ranks suspicious regions, estimates N:C ratio and nuclear geometry, and generates a numerical report that links each class score to its spatial evidence.

**Fig. 1.**
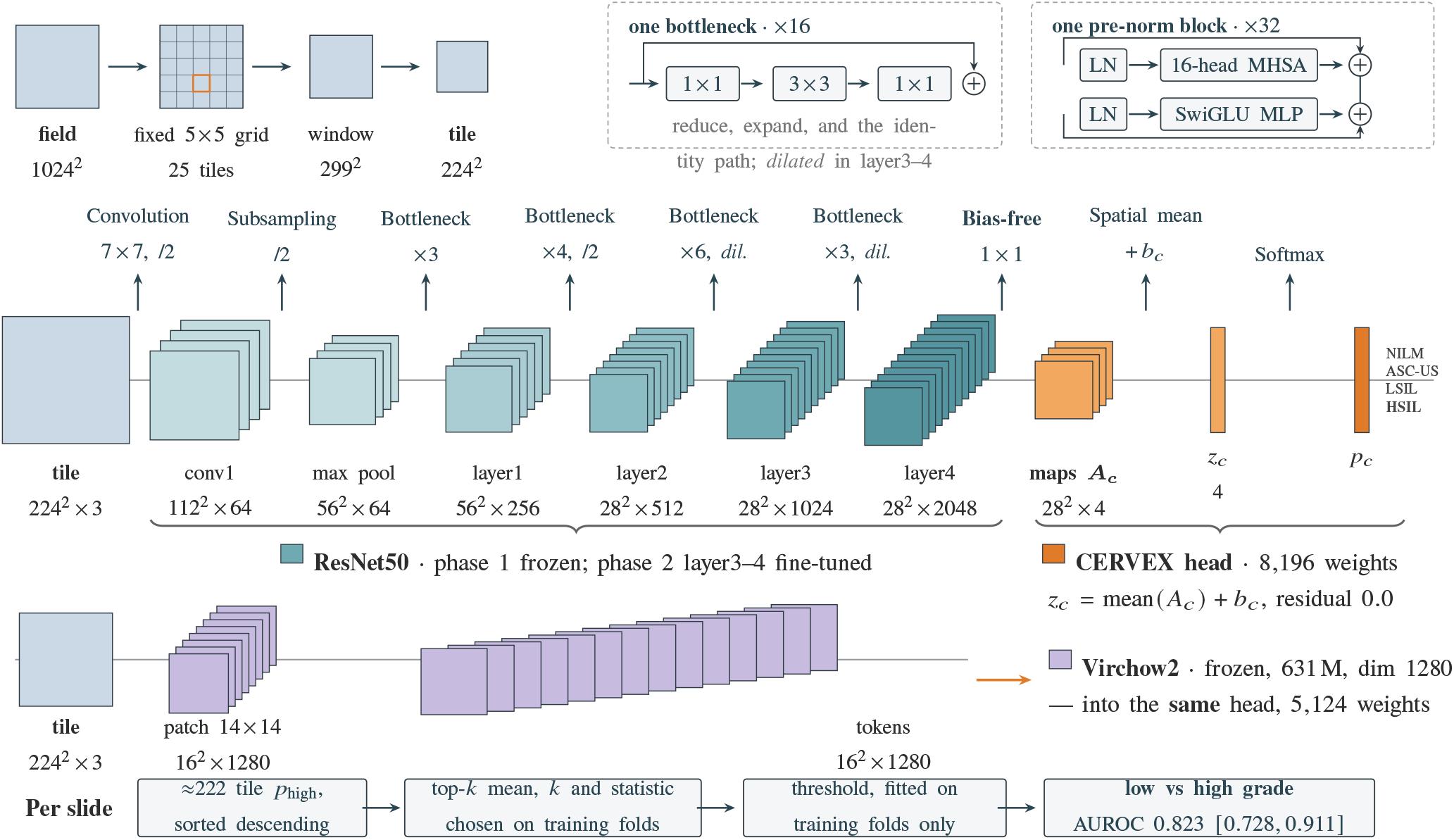
CERVEX processes one tile at a time. The ResNet50 readout is first trained with the trunk frozen; layer3 and layer4 are then fine-tuned at a lower learning rate. Virchow2 remains frozen. The orange readout converts the final spatial features into four evidence maps, whose means produce the four class scores (Eq. 1). Tensor labels give the exact dimensions; visual stack depth is log-scaled so large channel counts remain legible.

**Fig. 2.**
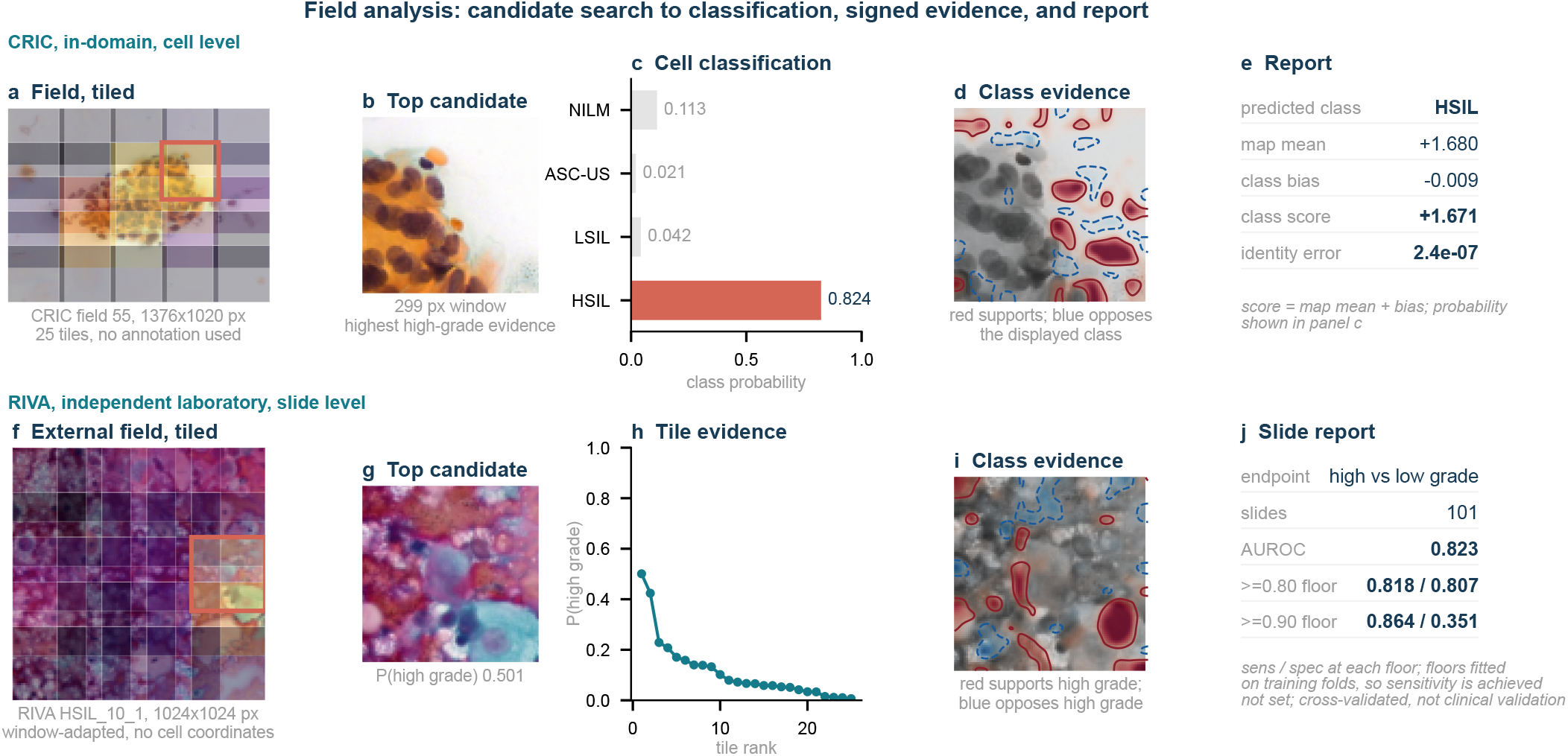
The same pipeline on real recorded runs, which Fig. 1 gives only as structure. *Top, CRIC, in-domain:* (a)–(e) a field on the fixed grid, the strongest high-grade tile, its posterior, its class-evidence map, and the report. *Bottom, RIVA, independent laboratory:* (f)–(j) the same chain with no annotation at inference, ending in the decision over 101 graded slides. Its crop window was chosen on RIVA, so this row is window-adapted rather than zero-shot. The two rows are separate cohorts, not spliced, and are illustrative. Red map values increase the displayed class score, whereas blue values decrease it. These values are signed score contributions, not segmented cells or verified abnormal-cell boundaries. Panel (e) exposes the exact calculation linking the map to the numerical class score.

Our experiments evaluate four contributions. First, we show that cell-level splitting inflates CRIC accuracy by 6.76 points and establish field blocking as the comparison basis (Section IV-A). Second, we compare CERVEX with a conventional dense classifier and test its evidence maps against independent location controls. Third, we test contour-derived concepts against an explicit non-morphological shortcut gate (Section IV-C). Finally, we perform fixed-grid field analysis without cell coordinates at inference (Section IV-E).

Our pipeline follows three linked steps. First, CERVEX classifies each cell. Next, it displays the class-specific evidence map that directly produced the score. Finally, it generates a numerical report that combines class probabilities, evidence strength, candidate-region ranking, and contour-derived morphology. Thus, the report summarizes the same spatial evidence and measurable cell features for review rather than generating a separate explanation after the prediction.

## II. Related Work

### A. Black-Box Medical AI and Post-Hoc Explanation

Medical-image classifiers can achieve high accuracy while revealing little about how they reached a decision. This black-box behaviour prevents clinicians from checking whether a model used relevant cell morphology or an irrelevant shortcut such as stain or scanner appearance [10], [11], [13]. Researchers often apply Grad-CAM, integrated gradients, SHAP, or LIME after training to visualize image importance [14]– [17]. However, these post-hoc maps estimate the model’s reasoning after it has already produced the class score; the classifier does not use the displayed map to make that score.

### B. Unfaithfulness in Post-Hoc Methods

Post-hoc explanations can look medically plausible without faithfully representing the model’s computation. Some maps remain similar after researchers randomize model parameters or labels [18], while perturbation scores change with the chosen masking procedure [19]. Therefore, visual appearance alone cannot establish faithfulness. CERVEX addresses this limitation by generating each class-evidence map before spatial averaging and computing the class score directly from that map (Eq. 1). The heatmap and numerical score consequently describe the same calculation rather than two separate procedures. We then test localization and morphology independently, because computational faithfulness alone does not prove medical relevance.

## III. METHODS

### A. Data, Endpoint, and Separation

CRIC provides 400 conventional Pap-smear fields from 118 patients and 11,534 annotated cells with nucleus-centre coordinates [20]. We use four Bethesda classes: NILM (6,779), ASC-US (606), LSIL (1,360), and a high-grade class that combines ASC-H, HSIL, and SCC (2,789) [21]. We extract a 224 × 224 crop around each marked cell and preserve the native image resolution.

We split complete fields into 280 fields/8,053 cells for training, 60/1,760 for validation, and 60/1,721 for testing. This field-blocked split prevents cells from the same image from crossing partitions. CRIC identifies each field but does not provide a patient identifier for every cell, so we cannot guarantee patient-level separation. We resample complete fields when estimating uncertainty [22]. We use RIVA to test transfer and fixed-grid analysis on microscope fields from another laboratory [23]. We use SIPaKMeD’s 4,049 contoured cell images to evaluate nucleus, complete-cell, and morphology measurements because CRIC does not provide contours [24].

### B. Class-Evidence Readout

CERVEX receives a 224 × 224 RGB cell image. Its primary ResNet50 encoder converts the image into 2,048 feature channels on a 28 × 28 spatial grid [25]. We use dilated convolutions in the final encoder stages to preserve this grid instead of reducing it to 7 × 7. A bias-free 1 × 1 convolution then converts the features into four 28 × 28 evidence maps, one each for NILM, ASC-US, LSIL, and HSIL. CERVEX averages each map, adds a class bias, and applies softmax to produce the four class probabilities. The foundation-model variants replace ResNet50 with Virchow2, UNI2, or H-optimus-0 while retaining the same additive evidence readout.

For microscope-field analysis, CERVEX applies the classifier to windows from a fixed grid. It ranks the windows by their class probabilities, displays the strongest candidate regions and evidence maps, and combines the results with contour-derived measurements in the final numerical report. Thus, one architecture links field search, cell classification, spatial evidence, and morphology reporting.

We compare CERVEX with a conventional classifier that uses the same ResNet50 encoder. The conventional classifier averages all spatial features into one vector before predicting the four classes, which removes location from the final decision. CERVEX instead keeps the spatial grid until it creates the four evidence maps. Therefore, the displayed map contains the spatial evidence that directly produces each score.

At image location (*i, j*), CERVEX combines the encoder channels *F_k_* with class-specific weights *w_ck_* to create evidence map *A_c_*. It averages that map and adds class bias *b_c_* to obtain score *z_c_*:

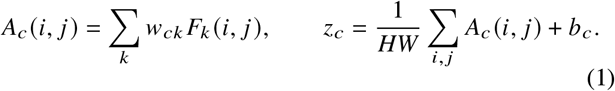

Softmax converts the four scores into class probabilities. Positive map values increase a class score, whereas negative values decrease it. This readout follows classical CAM [12]. We verify the map-to-score equality numerically for every model. To show why CERVEX selected class *p* instead of alternative *q*, we subtract their maps: *C_p,q_* = *A_p_* − *A_q_*. Positive regions support *p*, and negative regions support *q*.

We train each classifier from three random initializations. We give rare classes larger loss weights and randomly rotate and flip training images. First, we train the readout with the encoder frozen. Next, we fine-tune the final encoder stages at a lower learning rate. We select the checkpoint with the lowest validation loss. Finally, we compare both classifiers on the same cells, resample complete fields 10,000 times for uncertainty, and report each class’s sensitivity so that the large NILM class cannot hide errors in rarer classes.

### C. Spatial Evidence Validation and Shortcut Auditing

We perform two tests. First, we test whether the evidence map points to the annotated cell. We move the target cell away from the image centre in 168 crops and record whether the brightest map location falls within 8, 16, or 24 pixels of its nucleus. We compare CERVEX with Grad-CAM, a random map, and a map that always points to the crop centre. The off-centre design prevents a method from succeeding simply by highlighting the middle of every image. We also delete and insert highlighted regions and randomize model weights to test whether the map responds to changes in the input and model [18], [19].

Second, we test whether colour alone can predict the cell label without using spatial morphology. We train a simple classifier on global colour summaries and compare its score with scores from labels shuffled within each field. Shuffling within a field holds the shared imaging conditions constant. We repeat the test after Reinhard normalization [26] and after replacing each colour channel with within-image ranks. This audit identifies an available colour shortcut; it cannot determine whether that colour reflects biology or image acquisition.

CERVEX addresses these problems at two levels. First, it computes every class score directly from its evidence map, so the model cannot produce the score without also producing the corresponding spatial explanation. Unlike Grad-CAM and other post-hoc methods, CERVEX needs no separate backward-pass approximation. Second, the off-centre controls and colour-only probes test whether the apparent explanation comes from meaningful spatial evidence, a centre bias, or an easier colour shortcut. This design makes the heatmap, class score, and numerical report easier to trace and audit. The shortcut test detects bias; it does not claim to remove every source of bias.

### D. Creating the Numerical Cell Report

CERVEX first separates the nucleus and cytoplasm because these two regions define most of the reported cell measurements. CRIC and RIVA do not provide cell outlines, so we train this part on SIPaKMeD, which provides a contour around each nucleus and complete cell. A ResNet18 segmentation model receives a cell image and draws two masks: one for the nucleus and one for the complete cell.

CERVEX calculates the numerical report directly from these masks. It measures the N:C ratio, relative nuclear size, elongation, circularity, compactness, nucleus position, and perinuclear clearing. Figure 3c defines these quantities. The images do not provide a micrometre scale, so CERVEX reports relative measurements rather than physical size. These measurements describe visible cell structure; they do not act as fixed rules that assign a Bethesda class.

**Fig. 3.**
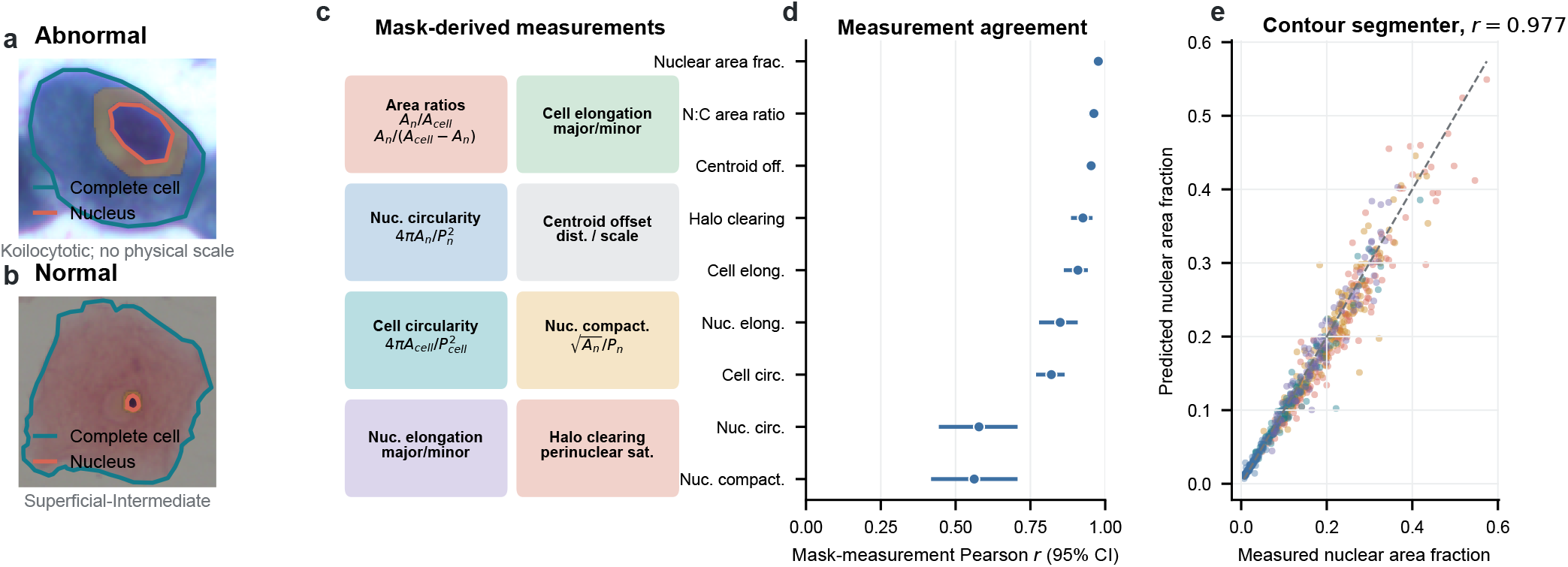
SIPaKMeD contour-grounded measurements. (a, b) Test crops with published contours. (c) Nine mask-derived quantities. (d) Correlation between measurements from predicted and reference rasterized masks, with 95% grouped-bootstrap intervals. (e) Predicted-mask versus reference-mask nuclear area fraction (*r* = 0.977 [0.970, 0.983], *n* = 642). Shape indices use standardized image coordinates.

We check the report in two steps. First, Dice overlap tests how closely the predicted masks match the supplied contours. Second, error and correlation test how closely each reported number matches the value calculated from those contours. We train three models, select them using validation data, and evaluate them on held-out cells. We also compare models that use RGB or reconstruction-error features with a colour-and-crop control. This final comparison tests whether the report captures cell morphology beyond simple image colour and framing [27], [28]. Clinical use would additionally require microscope calibration and reader-agreement testing [29].

## IV. Results

### A. Classification Accuracy and Data Splitting

We first tested whether the data split changes the reported accuracy. A cell-level split places cells from the same microscope field in both training and evaluation data: 322 of 340 fields were shared. This raised accuracy from 0.847 with field blocking to 0.915, an increase of 6.76 points. Macro-F1 rose from 0.708 to 0.835. The split therefore changed performance more than any architecture change in this study.

We next compared CERVEX with a conventional dense classifier on the separate held-out partition. CERVEX reached 0.776 accuracy and the dense classifier reached 0.780 across 1,721 cells from 60 fields. The difference was −0.45 points (95% CI [−1.70, +0.76], *p* = 0.475), so this experiment did not resolve an accuracy difference.

Table I summarizes the remaining comparisons. On validation data, the readout difference across ten seeds was +0.0034 [−0.0025, +0.0095]. Replacing ImageNet ResNet50 features with three frozen pathology backbones and combining their predictions increased accuracy from 0.851 to 0.902 (*p* = 0.0050). Every CERVEX variant retained an exact map-to-score identity.

**TABLE I.** Four-class classification results and whether each model provides an exact class-evidence map.

| Configuration | Accuracy | $\Delta$ vs. block reference [95% CI] | $p$ | Map identity error |
| --- | --- | --- | --- | --- |
| *(a) Readout contrast, ImageNet trunk, ten seeds* |  |  |  |  |
| Dense head, no evidence map | 0.833 | reference | — | none |
| **CERVEX readout** | **0.836** | +0.003 [−0.003, +0.010] | 0.269 | **0.0** |
| *(b) Split protocol, CERVEX dilated arm, three seeds* |  |  |  |  |
| Field-blocked, used throughout | 0.847 | reference | — | 0.0 |
| Cell-level shuffle, 322/340 fields shared | 0.915 | +0.068, a split artifact | — | 0.0 |
| *(c) Frozen pathology trunk, CERVEX readout refit* |  |  |  |  |
| ResNet50, ImageNet | 0.851 | reference | — | 0.0 |
| H-optimus-0 | 0.881 | +0.030 [−0.009, +0.069] | 0.1316 | 0.0 |
| UNI2 | 0.886 | +0.036 [+0.003, +0.072] | 0.0368 | 0.0 |
| Virchow2 | 0.895 | +0.044 [+0.009, +0.084] | 0.0128 | 0.0 |
| **Three-trunk ensemble + rotation TTA** | **0.902** | +0.051 [+0.016, +0.091] | 0.0050 | **0.0** |
| *(d) External laboratory, no target-label training, 16,698 cells, 51 slides, four readers* |  |  |  |  |
| Cell-level accuracy, per reader | 0.659–0.704 | balanced 0.668–0.735 | — | 0.0 |

Blocks (a)–(c) share the field-blocked CRIC validation partition (1,760 cells, 60 fields); (d) uses RIVA [23]. Within each block, $\Delta$ compares a row with that block’s reference using a paired grouped bootstrap. Residual is the largest observed gap between a class logit and its evidence-map mean plus bias; the dense head has no such map.

For abnormal-cell detection, sensitivity and specificity are more informative than overall accuracy. At a validation sensitivity target of at least 90%, CERVEX reached specificity 0.918 [0.874, 0.952], compared with 0.886 for the dense classifier (Table V). These are cell-level results, not patient-level screening results.

Table II shows that sensitivity increased as abnormality became more severe. Most errors therefore occurred in the borderline categories. Rejecting low-confidence predictions raised retained-set accuracy to 0.975 at 50% coverage, but it did so by deferring difficult cases, including 72.1% of LSIL cells.

**TABLE II.**
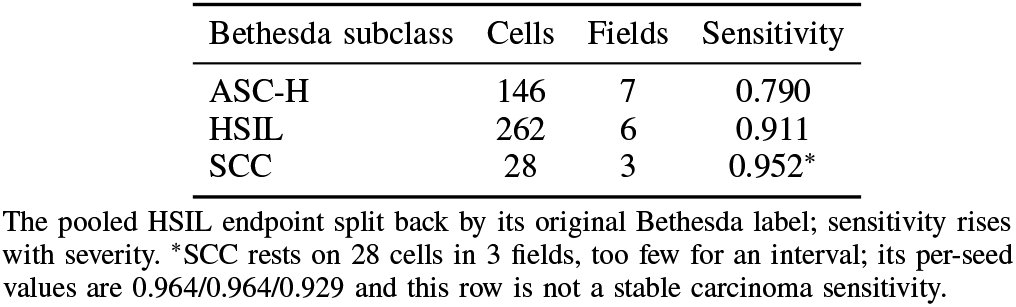
Where the errors fall on the field-blocked CRIC validation partition.

| Bethesda subclass | Cells | Fields | Sensitivity |
| --- | --- | --- | --- |
| ASC-H | 146 | 7 | 0.790 |
| HSIL | 262 | 6 | 0.911 |
| SCC | 28 | 3 | 0.952* |

The pooled HSIL endpoint split back by its original Bethesda label; sensitivity rises with severity. \*SCC rests on 28 cells in 3 fields, too few for an interval; its per-seed values are 0.964/0.964/0.929 and this row is not a stable carcinoma sensitivity.

**TABLE II.**
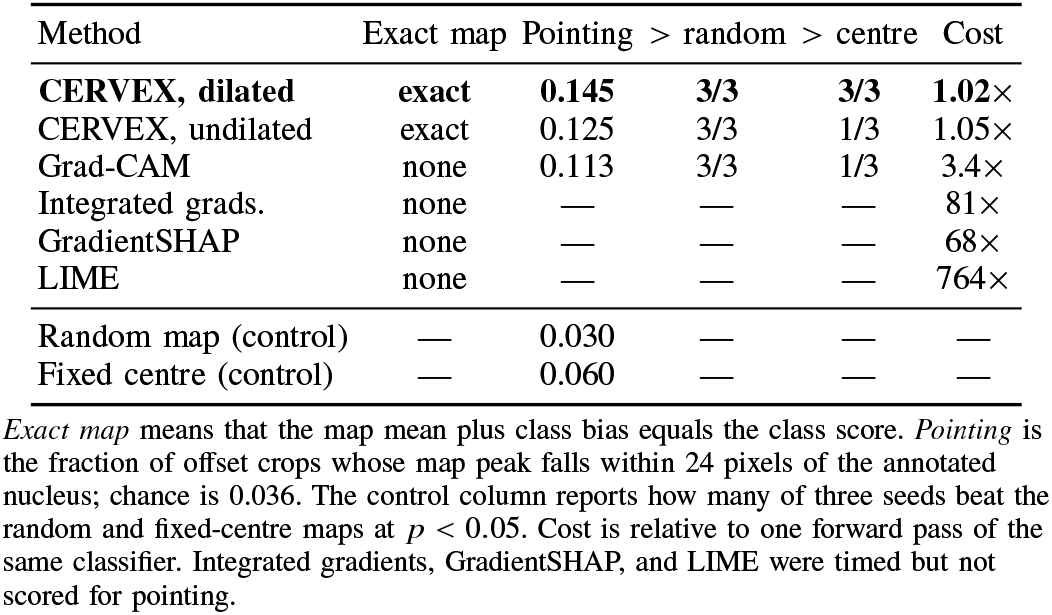
Explanation methods on the same CRIC crops and the same backbone.

### B. Does the Evidence Map Point to the Cell?

We moved the target cell away from the crop centre to prevent a centre-fixed heatmap from passing automatically. Across 168 crops, the dilated CERVEX map placed its peak within 24 pixels of the annotated nucleus in 0.145 of cases. This was 4.1 times the random rate of 0.036, and it beat both the random and fixed-centre controls in all three seeds (*p* ≤ 0.0106).

Grad-CAM reached 0.113 pointing accuracy, but it beat the fixed-centre control in only 1/3 of three seeds. The direct CERVEX-versus-Grad-CAM comparison did not resolve a difference (*p* = 0.113). Thus, the experiment supports localization beyond the two simple controls, but does not establish that CERVEX localizes better than Grad-CAM.

CERVEX creates its map during the same forward pass as the class score, adding 1.9% runtime. Grad-CAM required 3.4× one forward pass, integrated gradients 81×, and LIME 764×. Deletion and insertion results changed direction across seeds, so we treat them as diagnostics rather than evidence that one method is better.

### C. Are the Numerical Measurements Reliable?

Before evaluating morphology, we tested whether colour alone could predict the label. A classifier using only global colour summaries reached AUROC 0.8996 on CRIC. Rank transformation reduced this colour-only result to 0.4840, while a full image classifier still reached 0.992 [25]. Colour therefore provides an available shortcut, although this test cannot determine whether the colour comes from biology or image acquisition. The within-field test reached median 0.9697, compared with 0.7500 after label shuffling, and was higher in 88.6% of 219 fields (*p* = 2.86 × 10^−34^).

We then tested the segmentation and measurement pipeline on 642 SIPaKMeD cells. Nucleus and complete-cell Dice reached 0.932 and 0.942. Five measurements exceeded correlation *r* = 0.90 with the contour references: relative nuclear area (0.977), N:C ratio (0.962), nucleus position (0.953), perinuclear clearing (0.926), and cell elongation (0.909). Cell circularity and nucleus elongation reached 0.820 and 0.850. Nucleus circularity and compactness were weaker at 0.578 and 0.562 (Fig. 3). Correlation measures agreement in ordering; it does not define a clinical cutoff.

Accurate measurements do not automatically mean that the classifier uses them. A control using only colour and coarse crop geometry reached 0.847 balanced accuracy. The best morphology-concept classifier reached 0.875 ± 0.022, and the difference from the control was not resolved (*p* = 0.068). Ordinary image features also ranked above predictive-error features for every tested readout. In this experiment, predictive error did not improve concept-based classification beyond the shortcut control.

### D. Testing Stronger Encoders and Another Laboratory

We first replaced ResNet50 with three models trained previously on large pathology image collections. Accuracy reached 0.8949 with Virchow2, 0.8864 with UNI2, and 0.8807 with H-optimus-0 (Table Ic). All three kept the map identity error at 0.0. CERVEX can therefore use a stronger encoder without breaking the direct link between its heatmap and class score.

We next applied the CRIC-trained model to RIVA images from another laboratory, without training on RIVA labels. On 2,425 cells marked by multiple readers, AUROC was 0.828 [0.795, 0.858], with sensitivity 0.818 and specificity 0.686. These values were lower than the CRIC operating point, showing that a model trained in one laboratory does not transfer unchanged to another. Candidate localization remained better than random, but its top-ten precision ranged from 0.136 to 0.312 depending on which reader’s marks were used. The readers differed by 7.7× in how many cells they marked.

Finally, we retrained only the CERVEX readout using labelled RIVA cells. Training with a balanced mixture of CRIC and RIVA examples produced AUROC 0.911 (SD 0.009), sensitivity 0.869, and specificity 0.796 (Table IV). RIVA labels improved AUROC and specificity over CRIC-only training. Combining both datasets improved specificity over RIVA-only training, but did not resolve a further AUROC improvement. In simple terms, local training helped CERVEX reject normal cells from the new laboratory more reliably.

**TABLE IV.** How the training data changed cell classification on RIVA.

| Training data | AUROC | Sens. | Spec. |
| --- | --- | --- | --- |
| Original CRIC model, two scales | 0.845 | 0.874 | 0.598 |
| CRIC only, matched setup | 0.861 | 0.864 | 0.671 |
| RIVA only | 0.896 | 0.844 | 0.723 |
| **CRIC + RIVA, balanced** | **0.911** | **0.869** | **0.796** |

Results use repeated slide-grouped cross-validation. The first two rows use no RIVA labels, although RIVA was used to choose the wider image scale. The last two rows use labelled RIVA cells for training.

Rank normalization did not solve the transfer gap. It reduced CRIC accuracy from 0.8938 to 0.8653 and RIVA top-ten precision from 0.300 to 0.178. Detecting a colour shortcut and generalizing to another laboratory are therefore separate problems.

### E. Combining Field Results into One Slide Result

The final experiment asked whether CERVEX could combine many field predictions into one slide result. RIVA contains 57 low-grade slides and 44 high-grade slides, for 101 slides in total.

We tested two approaches. The first used cells already marked by a reader and reached AUROC 0.755 [0.644, 0.840]. The second searched every supplied field with a fixed grid and required no cell coordinates at inference. It reached AUROC 0.823 [0.728, 0.911] across 22,375 image regions. The uncertainty ranges overlap, so these results do not show that one approach is better.

This result supports automatic analysis after a microscope field has been captured. It does not yet test whole-slide screening: RIVA supplies selected fields rather than complete digital slides and contains only one normal slide. Table V therefore shows several operating points instead of presenting one threshold as clinically fixed.

**TABLE V.**
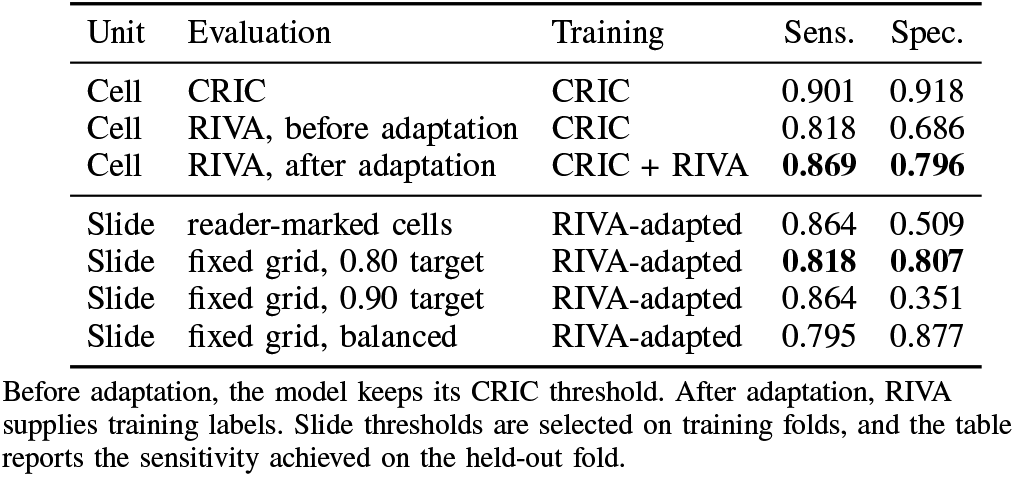
Sensitivity and specificity for cell detection and slide classification.

| Unit | Evaluation | Training | Sens. | Spec. |
| --- | --- | --- | --- | --- |
| Cell | CRIC | CRIC | 0.901 | 0.918 |
| Cell | RIVA, before adaptation | CRIC | 0.818 | 0.686 |
| Cell | RIVA, after adaptation | CRIC + RIVA | **0.869** | **0.796** |
| Slide | reader-marked cells | RIVA-adapted | 0.864 | 0.509 |
| Slide | fixed grid, 0.80 target | RIVA-adapted | **0.818** | **0.807** |
| Slide | fixed grid, 0.90 target | RIVA-adapted | 0.864 | 0.351 |
| Slide | fixed grid, balanced | RIVA-adapted | 0.795 | 0.877 |

Before adaptation, the model keeps its CRIC threshold. After adaptation, RIVA supplies training labels. Slide thresholds are selected on training folds, and the table reports the sensitivity achieved on the held-out fold.

## V. Discussion

CERVEX addresses the black-box problem by linking every class score to a class-specific evidence map. The separate held-out comparison did not resolve an accuracy difference between CERVEX and the conventional dense classifier. However, only CERVEX produced an evidence map whose mean plus bias exactly reproduced the class score, with identity error 0.0 across every tested encoder, seed, and cohort. Generating this map added 1.9% runtime. Its peak also reached the annotated nucleus 0.145 of the time, compared with a chance rate of 0.036, and beat both location controls in all three seeds. These findings show that the displayed heatmap belongs to the model’s calculation and usually points toward the target cell. They do not show that every highlighted region represents a medically abnormal structure.

The numerical report adds a separate and testable description of cell morphology. Segmentation reached Dice 0.932 for the nucleus and 0.942 for the complete cell. Relative nuclear area, N:C ratio, nucleus position, perinuclear clearing, and cell elongation each exceeded correlation *r* = 0.90 with measurements from the supplied contours. Nuclear boundary measurements were less reliable. The morphology-concept classifier also did not resolve an improvement over the colour-and-crop control, and predictive-error features did not improve this comparison. Therefore, the report can present measured image features for review, but the present study does not claim that these measurements alone explain the Bethesda prediction or define clinical thresholds.

The RIVA experiments show why local evaluation matters. The CRIC-trained model lost performance when applied directly to images from another laboratory. Retraining the readout with labelled RIVA examples raised abnormal-cell AUROC to 0.911, with sensitivity 0.869 and specificity 0.796. Fixed-grid field analysis then separated low-from high-grade abnormal slides with AUROC 0.823 [0.728, 0.911], without cell coordinates at inference. These results support automatic analysis within captured microscope fields and show that a new laboratory may require local readout training and threshold selection.

The study still has defined boundaries. Field blocking prevents cells from the same image from crossing partitions, but it cannot guarantee patient independence for records that lack verified patient identifiers. The SCC estimate uses few cells, SIPaKMeD retains acquisition cues, and nucleus-centre pointing is less specific than comparison with expert structure outlines. RIVA supplies selected microscope fields and only one normal slide, so it cannot test complete-slide screening. Future validation should use calibrated image scale, blinded expert annotations, complete digital slides, and multiple laboratories before CERVEX supports clinical decision making [13].

### Future Work

CERVEX will next be tested on complete digital slides and against blinded expert markings. A public research interface is being prepared to display class scores, evidence maps, and candidate regions from captured microscope fields.

## VI. Conclusion

Cell-level splitting inflates CRIC accuracy by 6.76 points, showing why the split unit must be reported. CERVEX makes each class score exactly traceable to its map with residual 0.0 across every tested trunk, seed, and cohort, while adding 1.9% to one forward pass. Its readout differs from a dense head by +0.003 accuracy points, within this design’s resolution. These results support separate reporting of computation, anatomy, concepts, and shortcut resistance. Expert structure annotations and measurement agreement are the next requirements for pathology-facing interpretation.

## Data Availability

All image data analyzed in this study are publicly available. CRIC is available through Figshare at https://doi.org/10.6084/m9.figshare.c.4960286.v2. RIVA is available through Zenodo at https://doi.org/10.5281/zenodo.17288879. SIPaKMeD is available from the University of Ioannina at https://www.cs.uoi.gr/~marina/sipakmed.html. This study generated no new human participant data.

https://doi.org/10.5281/zenodo.17288879

https://www.cs.uoi.gr/~marina/sipakmed.html

## Ethics, Data, and Conflicts of Interest

This secondary analysis used public, de-identified data; no approval or consent was required. The authors declare no conflicts.

## Acknowledgment

The authors thank Bangkok Christian College, Mahidol University, and Chulalongkorn University. Language models assisted English editing; the authors reviewed and remain responsible for all content.

